# Pholcodine consumption increases the risk of perioperative anaphylaxis to neuromuscular blocking agents: the ALPHO case-control study

**DOI:** 10.1101/2022.12.12.22283353

**Authors:** Paul Michel Mertes, Nadine Petitpain, Charles Tacquard, Marion Delpuech, Cédric Baumann, Jean Marc Malinovsky, Dan Longrois, Aurélie Gouel-Cheron, Diane Le Quang, Pascal Demoly, Jean Louis Guéant, Pierre Gillet

**Author notes:** **Corresponding authors** Paul Michel MERTES, MD, PhD, Department of Anesthesia and Intensive Care, Hôpitaux Universitaires de Strasbourg, Nouvel Hôpital Civil, 1 Place de l’Hôpital - BP 426 67091 STRASBOURG CEDEX - France, Work. **Contribution of authors** PMM, NP, JLG, CB and PG designed and planned the study. PMM, NP, DL, JMM and PD implemented the study. CB, MD, PMM, NP and CT accessed and verified the data. PMM, NP, CT, AG, DLQ, JLG, CB, MD and PG analysed and interpreted the trial data. PMM, NP and CT wrote the first draft of the manuscript. All authors read and approved the final version of the manuscript, had full access to all the data in the study, and had final responsibility for the decision to submit for publication. All the members of the ALPHO study group were in charge of inclusion of cases, and controls, and data collection and approved the final version of the manuscript.

## Abstract

**Background:** Neuromuscular blocking agents (NMBAs) are the leading cause of perioperative anaphylaxis (POA), most reactions being IgE mediated. Allergic sensitization induced by environmental exposure to other quaternary ammonium-containing compounds, such as pholcodine, has been suggested. The aim of this study was to assess the relationship between pholcodine exposure and NMBA-related POA.

**Methods:** ALPHO is a multicentre case-control study, comparing pholcodine exposure within a year before anaesthesia between patients with NMBA-related POA (cases) and control patients with uneventful anaesthesia. Each case was matched to two controls by age, sex, type of NMBA, geographic area, and season. Pholcodine exposure was assessed by a self-administered questionnaire and pharmaceutical history retrieved from pharmacy records. The diagnostic values of anti-pholcodine and anti-quaternary ammonium specific IgE (sIgE) were also evaluated.

**Results:** Overall, 167 cases were matched with 334 controls. NMBA-related POA was significantly associated with pholcodine consumption (OR =4.2; CI95% 2.3-7.0) and occupational exposure to quaternary ammoniums (OR = 6.1; CI95% 2.7-13.6). Anti-pholcodine and anti-quaternary ammonium sIgEs had a high negative predictive value (99.9%) but a very low positive predictive value (< 3%) for identifying NMBA-related POA.

**Conclusion:** Patients exposed to pholcodine 12 months prior to NMBA exposure have a significantly higher risk of a NMBA-related POA. The low positive predictive values of pholcodine and quaternary ammonium sIgEs precludes their use to identify a population with a high risk of NMBA-related POA. The strong association of NMBA-related POA with occupational exposure suggests that other environmental factors may also lead to sensitization to NMBAs.

## INTRODUCTION

Neuromuscular blocking agents (NMBAs) represent one of the leading causes of perioperative anaphylaxis (POA) ^1-5^. Although rare, with an estimated frequency of 184 reactions per million anaesthesia in France, NMBA-related POA are largely unpredictable, potentially catastrophic and remain a major concern for anaesthesia providers^1,3,6^.

The main mechanism involves the interaction between specific IgE and an epitope present in the structure of the NMBA resulting in massive activation of mast cells and basophils.^7^ Quaternary ammonium (QA) groups, essential for the relaxant properties of any NMBA, are considered the main part of the allergenic structures recognized by IgE antibodies detected in the sera of most patients who experience NMBA-related POA. Classically, this paradigm requires prior exposure to NMBAs to induce sensitization.

However, many subjects who react to a NMBA have never been exposed to one of these drugs, challenging the immunological dogma of the necessity of a previous exposure to elicit the production of specific IgEs.^8^ This led to the speculation that the origin of allergic sensitization could be induced by environmental exposure to other compounds containing a substituted ammonium ion.

In 2005, Florvaag and Johansson proposed pholcodine, an opiate that contains an ammonium group, as an important risk factor for NMBA sensitization, and gradually strengthened this hypothesis until 2011^9-13^. Pholcodine-containing drugs were withdrawn from the Norwegian market in March 2007, and a decrease in the prevalence of specific antibodies recognizing pholcodine but also the NMBA suxamethonium was observed in the general population over the subsequent two years^9^.

As a result, in 2011, the European Medicines Agency (EMA) launched a referral procedure for the re-evaluation of the benefit/risk ratio of pholcodine. The EMA decided to keep pholcodine on the European market but to request a case-control study to further investigate a possible increased risk of NMBA-related POA associated with pholcodine exposure. The objective of the ALPHO study was to investigate the relationship between exposure to pholcodine during the year preceding the general anaesthesia including a NMBA injection and the onset of a NMBA-related POA. The secondary objective focused on the diagnostic value of QA and PHO specific IgEs for the prediction of POA.

## METHODS

### Study design

The ALPHO study (NCT02250729) was a multicentre case-control study, supported by the EMA and funded by the pharmaceutical companies marketing pholcodine containing antitussive drugs. This study was conducted between 2014 and 2020 in 24 academic French anaesthesia departments and allergology units constituting to the GERAP network (Groupe d’Etude des Réactions Anaphylactiques Peropératoires)^1,14-16^.

### Patient screening and enrolment

Patients who experienced a POA involving an NMBA (cases) were identified by French anaesthetists and referred to one of the participating centres. Each reaction was reported to the ALPHO investigators and triggered the search for two matched controls who had been anesthetized with the same NMBA but had not experienced a POA. The matching criteria were: i) age (2-12, 12-17, 18-65, more than 65 years-old); ii) gender; iii) type of NMBA (suxamethonium; steroidal NMBA (rocuronium or vecuronium); benzylisoquinoline (atracurium, cisatracurium, or mivacurium); iv) geographic area (northern or southern France); and v) anaesthesia period corresponding to the date of POA +/-90 days.

Each case underwent NMBA allergy skin tests (prick tests (PT) and intradermal skin tests (IDT)) according to the EAACI guidelines^17^. A centralized retrospective confirmation of NMBA-related cases was performed by two specialists blinded for pholcodine exposure. Cases were excluded if they did not have at least one positive skin test for one of the injected NMBAs, or if their skin tests were not performed according to EAACI guidelines ^17^.

Controls were included according to matching criteria, by an anaesthetist investigator of the ALPHO study, during their hospital stay. They were required to complete at least the medication self-administered questionnaire. A history of POA during anaesthesia and pregnancy at the time of the inclusion were exclusion criteria.

The data collected included medical history, drugs administered during anaesthesia, characteristics and management of the POA, and profession. Patients reporting current or past cleaning profession or hairdressers were considered professionally exposed to QAs^18,19^. Blood tryptase and histamine measurements were performed according to French guidelines: at T0 (30 minutes after the reaction), T1 (1-2 hours after the reaction), and T2 (at least 24 hours after the reaction).

### Assessment and definition of pholcodine exposure

Exposure to pholcodine over the last 12 months before the anaesthesia index was assessed in cases and controls from two separate sources: 1) a self-administered questionnaire collecting the names and package visuals of all currently available pholcodine-containing drugs in France since 2014, as well as other marketed cough suppressants and non-pholcodine analgesics to serve as decoys. This questionnaire was completed during the allergy consultation for cases or during hospitalization for controls; 2) the patient’s pharmaceutical history retrieved from their community pharmacy records.

Patients were considered as exposed to pholcodine over the 12 months prior to anaesthesia if they reported taking at least one pholcodine-containing medication in the self-administered questionnaire, and/or if the medication history reported at least one dispensing of a pholcodine-containing medication.

### Pholcodine sensitization

Anti-pholcodine sIgE (PHO-sIgE) and anti-QA sIgE (QA-sIgE) were measured by two techniques: 1) ImmunoCAP® (Uppsala, Sweden) allergens c261 (pholcodine, PHO-c261) and c260 (QA-c260), and 2) Sepharose Ammonium Quaternaire-Fluorescence Immuno Assay (SAQ-FIA) with QA sIgE (QA-SAQ) results expressed by fixation rate (%) and PHO sIgE (PHO-SAQ) expressed by inhibition rate (%)^20^.

### Statistical analysis

Analyses were performed in SAS v9.4 (SAS Institute, NC, Cary, USA). Categorical variables were expressed as numbers and percentages and continuous variables as median and interquartile range. Comparison tests of proportions and means were performed using Chi2 test, Fisher’s exact test and Student’s t test or Man-Whitney test. For multivariate conditional logistic regression models, a bivariate analysis was performed to select the adjustment variables and only variables with a p-value ≤ 0.2 were candidates for multivariate conditional logistic regression models. The strength of association was estimated by adjusted ORs with their 95% confidence intervals (5%level of statistical significance of the two-tailed tests). Two multivariate models were compared 1) multivariate logistic regression model including all selected candidate variables; 2) stepwise candidate variable selection procedure (entry threshold at p = 0.2 and model exit threshold at 0.05). The model with the lowest AIC was retained for the results. Concordance analysis was based on Cohen’s Kappa coefficient.

## RESULTS

### Selection process

Between 21 July2014, and 20 July 2020, 937 patients were screened for eligibility (Figure 1). Of these patients, 282 experienced a POA possibly related to NMBAs and 655 had uneventful anaesthesia. Following centralized review blinded for pholcodine use, 78 patients with POA and 5 without POA were excluded. NMBA-related cases and controls were strictly comparable with respect to matching variables (Table S1). Twenty-seven cases could not be matched with controls. Finally, 167 NMBA-related POA cases were matched with 334 patients with uneventful anaesthesia.

**Figure 1:**
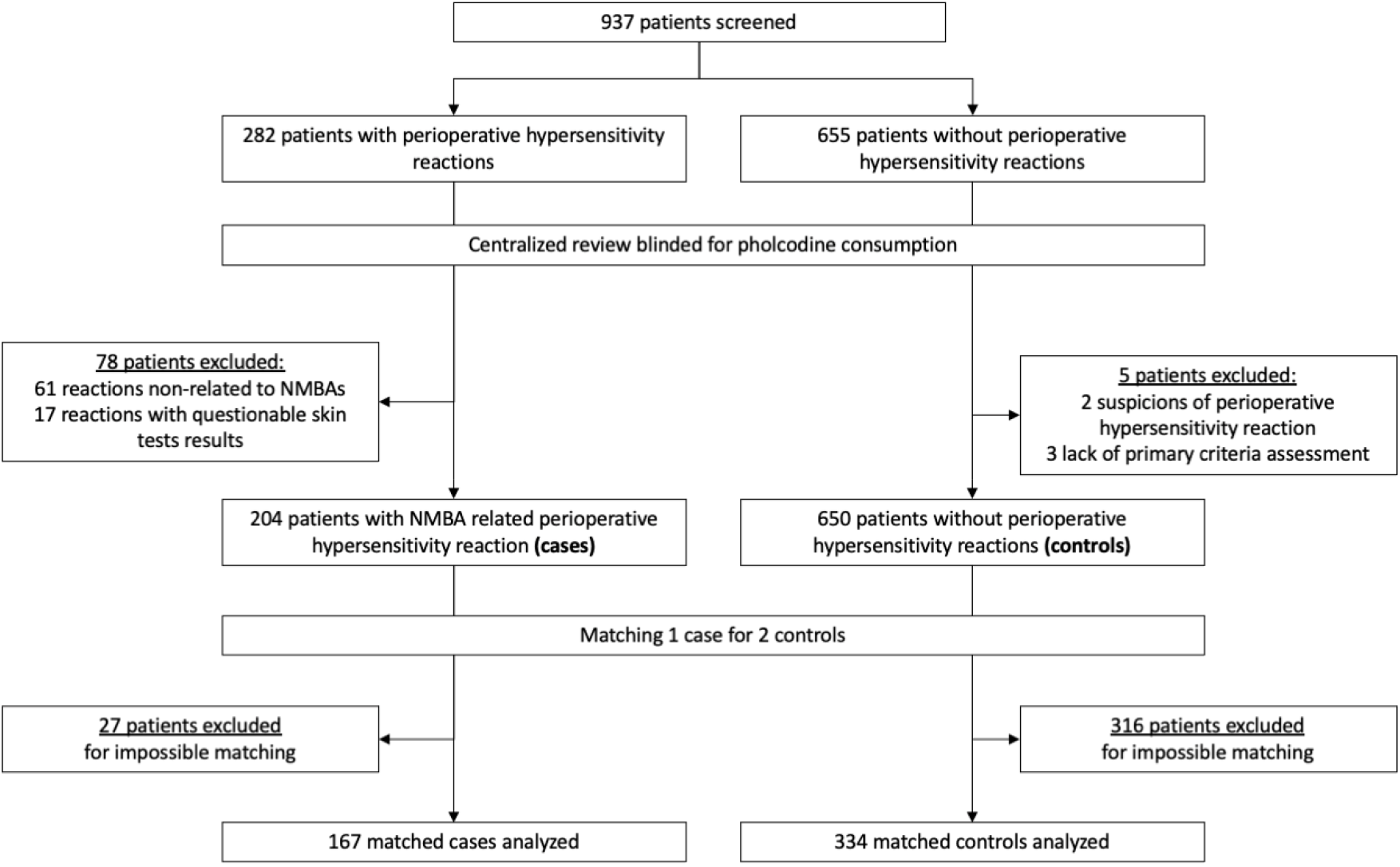
Flow chart of the ALPHO study. NMBA: neuromuscular blocking agents.

### Patients’ characteristics

The characteristics of the cases and controls are described in Table 1. Patients were predominantly female (cases 92 (55%) *vs*. controls 184 (55%)) and BMI was significantly higher in cases than in controls. No significant differences were observed regarding medical history or chronic medication. Cases reported significantly more atopy than controls. Table S2 shows the surgical procedure that led to the patient’s inclusion.

**Table 1:**
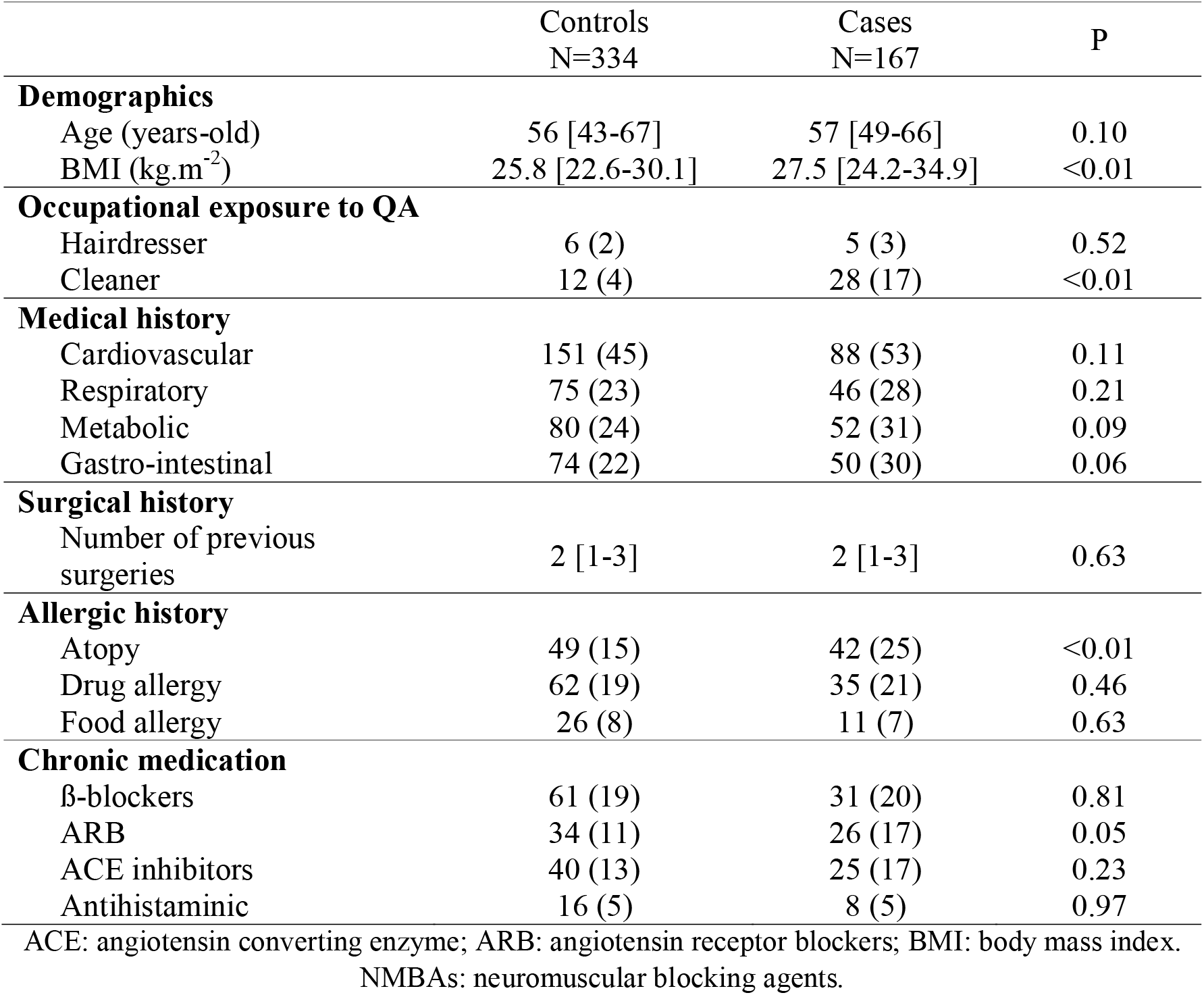
Characteristics of the case and control cohorts.

### Pholcodine consumption

The self-questionnaire was completed in 163 (98%) of cases and 333 (99%) of controls. Pharmacist-reported medication history was available in 121 (72%) cases and 172 (51%) controls. According to the self-questionnaire, 71 (43%) cases and 63 (19%) controls reported the use of pholcodine in the past year. When considering only the pharmacist reported medication history 26 (22%) cases and 7 (4%) controls ascertained the use of pholcodine. Eighteen (13%) cases and 3 (1%) controls reported the use of pholcodine in both sources. Overall, 79 (47%) cases and 67 (20%) controls reported the use of pholcodine in the year preceding the anaesthesia index (p<0.001).

### NMBA related perioperative hypersensitivity reaction

An NMBA-related POA occurred during an emergent surgery in 42 (25%) cases. In 20 (12%) cases, the patient was in the operating theatre for a bariatric surgery. Suxamethonium was used in 105 (63%) cases, rocuronium in 21 (13%) cases, atracurium in 53 (32%) cases, cisatracurium in 21 (13%) cases, and mivacurium in 3 (2%) cases (Table S3); patients may have received more than one NMBA.

Most of the POAs were severe with 14 (9%), 140 (84%), 13 (7%) reactions of grade II, III, and IV, respectively. The main clinical sign observed was cardiovascular collapse (n=134; 80%) (Table S3). Epinephrine was used in 145 (87%) cases to treat the POA.

The median tryptase level was 53 [18-113] µg/L at T0 (n=129; 77%), 33 [15-69] µg/L at T1 (n=114; 68%) and 5 [3-8] µg/L at the basal state (n=83; 50%). Figure 2 shows the distribution of tryptase according to the pholcodine consumption.

**Figure 2:**
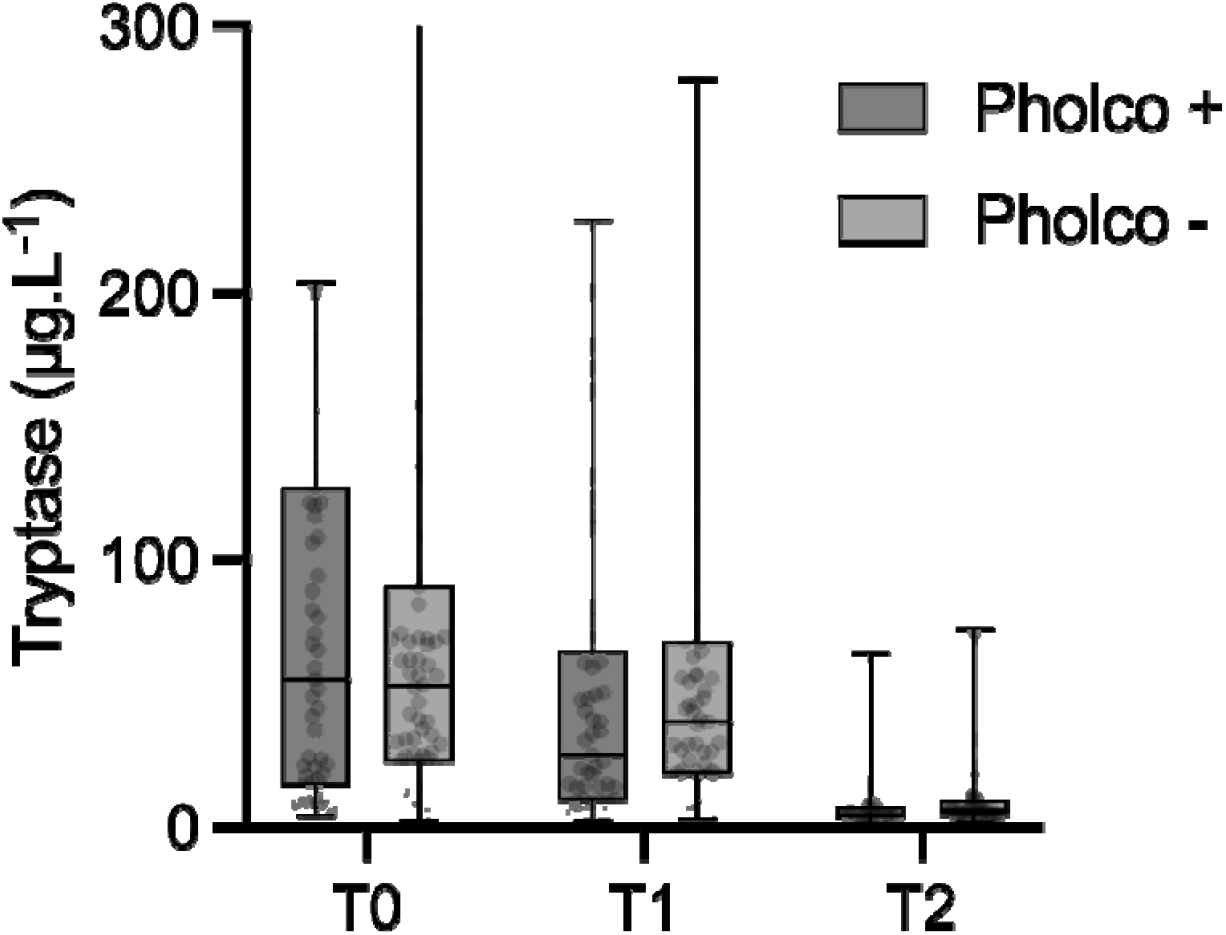
Serum tryptase level at T0 (<30 minutes), T1 (1-2 hours) and T2 (> 24 hours) after a neuromuscular blocking agent-related perioperative anaphylactic reaction, according to the consumption of pholcodine in the past year.

### Allergy workup

The allergy workup was performed within a median period of 9 [7-11] weeks following the reaction. All patients had positive skin tests to at least one injected NMBA. At the end of the allergic workup, suxamethonium was incriminated in 101 (60%) cases, rocuronium in 21 (13%) cases, atracurium in 35 (21%) cases, cisatracurium in 11 (7%) cases, and mivacurium in 2 (1%) cases. Three (2%) patients were sensitized to two NMBAs injected during the surgery (suxamethonium and atracurium).

### Multivariate analysis

The multivariable analysis showed that pholcodine consumption was associated with NMBA-related POA with an OR of 4.2 (CI95% 2.3-7.0) (Table 2). Occupational exposure to QA and hepato-gastro-intestinal history were also associated with NMBA-related POA with an OR of 6.1 (CI95% 2.7-13.6) and 2.1 (CI95% 1.3-3.3), respectively.

**Table 2:** Risk factors for neuromuscular blocking agents-related perioperative hypersensitivity reactions.

|  | Bivariate analysis |  | Multivariate analysis |  |
| --- | --- | --- | --- | --- |
|  | OR | p | OR | P |
| Pholcodine use | 4.5 (2.8-7.3) | <0.01 | 4.2 (2.5-7.0) | <0.01 |
| Occupational exposure to QA | 6.0 (2.9-12.8) | <0.01 | 6.1 (2.7-13.6) | <0.01 |
| Atopy | 1.8 (1.1-2.8) | 0.02 | - | - |
| BMI kg.m<sup>-2</sup> |  | 0.08 |  |  |
| BMI 25-30 kg.m<sup>-2</sup> | 1.1 (0.7-1.8) | 0.45 |  |  |
| BMI ≥ 30 kg.m<sup>-2</sup> | 1.7 (1.0-2.6) | 0.03 | - | - |
| Cardiovascular history | 1.4 (0.9-2.2) | 0.09 | - | - |
| Metabolic history | 1.5 (1.0-2.4) | 0.07 | - | - |
| Endocrine history | 1.8 (0.9-3.5) | 0.10 | - | - |
| Gastro-intestinal history | 1.5 (1.0-2.3) | 0.06 | 2.1 (1.3-3.3) | <0.01 |

Results of Odds Ratio (OR) are expressed as OR (95% confidence interval); QA, quaternary ammoniums; BMI, body mass index.

When considering only the self-questionnaire as the source of the pholcodine exposure, pholcodine consumption remained associated with NMBA-related POA with OR 4.1 (CI95% 2.4-6.8), as occupational exposure to QA (OR 6.3 (CI95% 2.8-14.4)) and hepato-gastro-intestinal history (OR 4.1 (CI95% 2.4-6.8)). When considering only the pharmacist-reported medication history, only pholcodine exposure (OR 4.8 (CI95% 1.6-14.7)) and occupational exposure to QA (OR 2.9 (CI95% 1.1-7.6)) remained significantly associated with NMBA-related POA. When considering pholcodine exposure only if both sources were positive, pholcodine exposure (OR 11.0 (CI95% 3.1-39.4)), occupational exposure to QA (OR 5.5 (CI95% 2.3-13.2)), hepato-gastro-intestinal history (OR 1.7 (CI95% 1.0-3.0)), and atopy (OR 2.1 (CI95% 1.1-3.9)) were significantly associated with NMBA-related POA.

### Specific IgEs

QA-SAQ sIgE were assayed in 160 (96%) cases and 334 (100%) controls and QA-c260 in 162 (97%) cases and 333 controls. PHO-SAQ sIgE were assayed in 160 (96%) cases and 334 (100%) controls and PHO-c261 in 162 (97%) cases and 333 controls. Table 3 shows the results of sIgE in cases and controls. Figure S1 shows the ROC curves for sIgEs. There was no difference for sIgE values between patients who reported using pholcodine in the previous year and those who did not, with the exception of PHO-c261 in controls (Figure 3). Both SAQ-FIA and ImmunoCAP® methods were well correlated: Pearson coeff (QA-SAQ/PHO-SAQ) 0.86, p<0.001; Pearson coeff. (QA-c260/PHO-c261) 0.85, p<0.001. Specific IgEs had a high negative predictive value of 99.9% but a very low positive predictive value of 2.8, 0.3, 5.3, and 0.4% for QA-SAQ, QA-c260, PHO-SAQ, and PHO-c261 respectively (Table 3). In the subgroup of cases and controls who reported previous-year pholcodine use, the presence of PHO-c261 sIgE in patients was the only factor significantly associated with NMBA-related POA with OR 60.7 (CI95% 25.6-143.8) (Table S4).

**Table 3:** Specific IgE levels in case and control patients from the ALPHO study and their respective diagnostic value.

|  | Controls | Cases | p | AUC | Optimal threshold | Se (%) | Sp (%) | PPV* | NPV* |
| --- | --- | --- | --- | --- | --- | --- | --- | --- | --- |
| QA-SAQ (%) | 1.0 [0.9-1.0] | 3.5 [2.8-5.5] | <0.001 | 0.98 | 1.8 | 94 (92-96) | 99 (99-100) | 2.8 | 99.9 |
| QA-c260 (kUI.L<sup>-1</sup>) | 0.0 [0.0-0.0] | 1.5 [0.3-5.6] | <0.001 | 0.95 | 0.04 | 87 (84-90) | 94 (92-96) | 0.3 | 99.9 |
| PHO-SAQ (%) | 0.0 [0.0-0.0] | 40.0 [23.0-45.0] | <0.001 | 0.96 | 10.0 | 92 (89-94) | 100 (99-100) | 5.3 | 99.9 |
| PHO-c261 (kUI.L<sup>-1</sup>) | 0.0 [0.0-0.0] | 2.2 [0.4-6.5] | <0.001 | 0.95 | 0.05 | 88 (88-94) | 96 (94-98) | 0.4 | 99.9 |

AUC: area under the curve; Se: sensitivity; Sp: specificity; PPV: positive predictive value; NPV: negative predictive value. \* Based on an estimated incidence of 184/1,000,000 anesthesia<sup>1</sup>

**Figure 3:**
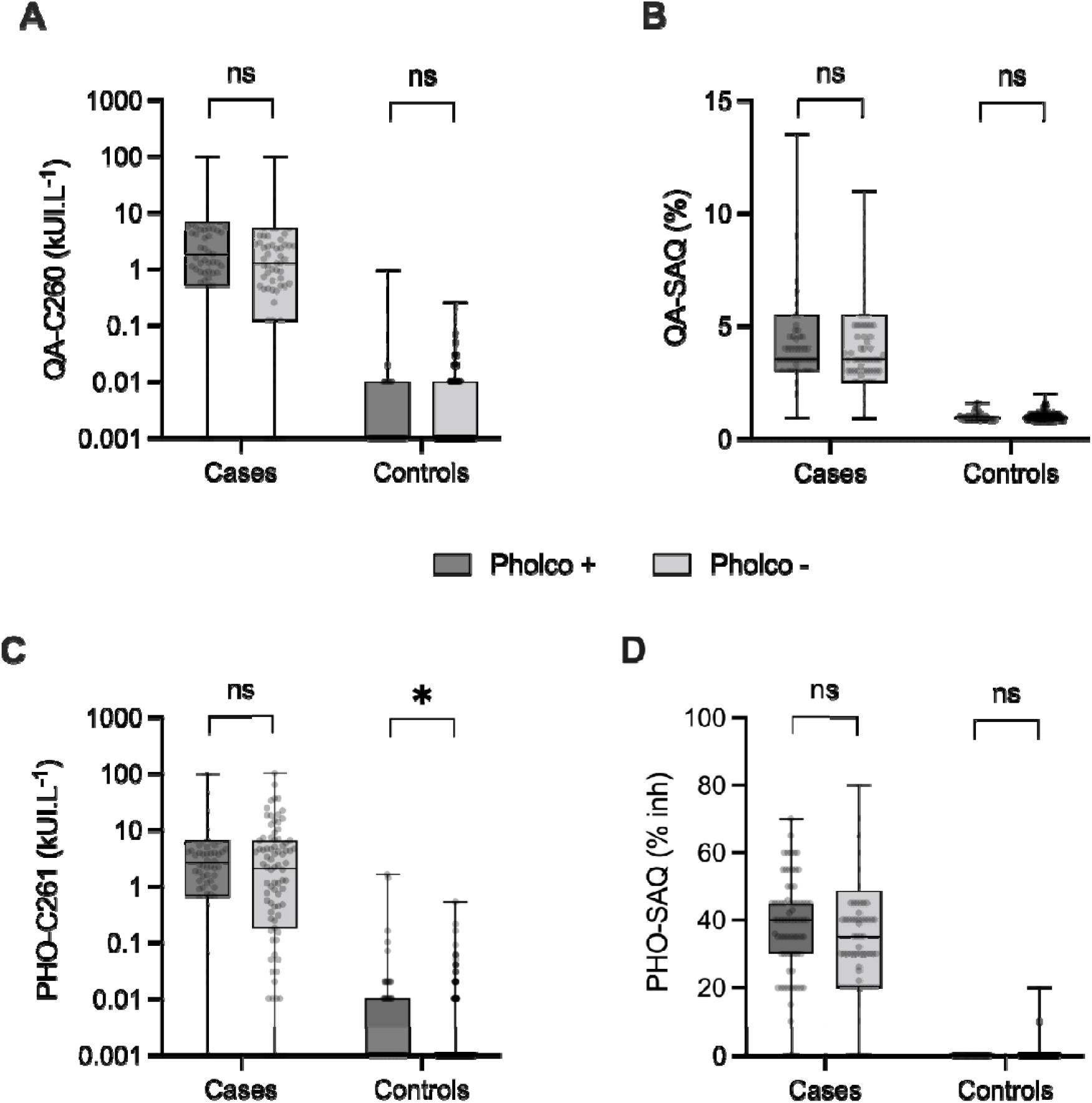
Specific IgE level in cases and controls according to their pholcodine consumption in the past year. Anti-quaternary ammoniums specific IgE (QA) using the Sepharose Ammonium Quaternaire-Fluorescence Immuno Assay (SAQ-FIA) method (QA-SAQ) or the ImmunoCAP® method (QA-c260). Anti-pholcodine specific IgE (PHO) using the SAQ-FIA method (PHO-SAQ) or the ImmunoCAP® method (PHO-c261). *p<0·05

## DISCUSSION

Patients exposed to pholcodine 12 months prior to anaesthesia have a significantly higher risk of a NMBA-related POA, which confirms the link between pholcodine exposure and NMBA-related POA. The strong association between occupational exposure and occurrence of NMBA-related POA suggests that other environmental factors may also lead to sensitization to NMBAs. QA-sIgE and PHO-sIgE have an excellent performance in discriminating cases from controls. Their negative predictive value is very high, indicating a low risk of NMBA-related POA if they are undetectable. However, the low positive predictive values of QA-sIgE and PHO-sIgE precludes their use to identify a population with a high risk of NMBA-related anaphylaxis.

The hypothesis of the role of pholcodine raised in 2005 by the Norwegian team of E. Florvaag ^9-13^, was based mainly on specific IgE tests, without any information on the confirmation of anaphylactic reactions to NMBAs by skin tests. In 2017, six years after the withdrawal of pholcodine in Norway, this team confirmed a decrease in sensitization to NMBAs in the general Norwegian population, especially in women aged 15 to 41 years^21,22^. At the same time, an observational study comparing pholcodine exposure in patients with POA related to NMBA versus patients who had POA related to cefazolin showed that pholcodine consumption was associated with a very significant increased risk of NMBA-related POA (OR=14.0, P<0.001)^23^. However, pholcodine exposure was estimated on the basis of information collected several years later by patient interview in only one third of the patients included.

Our study is the first fully case-control study comparing pholcodine exposure of patients with a confirmed POA to NMBA with controls anesthetized under the same conditions without POA. It clearly demonstrates that patients exposed to pholcodine 12 months before exposure to NMBA have a significantly higher risk of having an NMBA-related POA. This strong association is observed regardless of the source used to estimate pholcodine exposure, either by the self-questionnaire, the patient’s pharmaceutical history from his/her community pharmacist, or a combination of both, even when exposure was corroborated by the two sources.

Our results also suggest that, apart from pholcodine, other unidentified compounds containing tertiary or quaternary ammonium groups may act as sensitizing agents. These agents are widely present in the human environment among drugs, cosmetics, disinfectants, industrial materials, and foods. We observed that professional exposure to QA remained strongly associated with NMBA-related POA regardless of the sources of information. This possible role of environmental factors has long been suspected based on inhibition experiments of the binding of sIgE antibodies detected in patients suffering from POA by other compounds containing substituted ammonium ions epitopes^8,24,25^. This hypothesis has gained some support with the report of a high prevalence of sensitization to QA ions in several countries where pholcodine was not available ^10,26^, and with the demonstration that professional exposure to QA containing compounds among hairdresser students increases IgE-sensitization to NMBAs^18^.

In our series, a history of hepato-gastrointestinal disease was identified as an additional possible risk factor. This could be related to a higher probability of taking cough syrups (presence of cough in case of gastroesophageal reflux/hiatal hernia).

The results of QA-sIgE and PHO-sIgE showed an excellent performance in discriminating POA cases from controls, with a significant correlation between the results obtained with these different assays. This supports the recognition of common epitopes between the QA and PHO-sIgE tests, with a slightly better performance of the SAQ-FIA technique than the ImmunoCAP® assay.

The results of QA-sIgE and PHO-sIgE were not significantly influenced by pholcodine exposure, except for a statistical, although not biologically relevant, C261-PHO difference between controls. Thus, we were unable to detect a possible “booster effect” of pholcodine exposure on IgE production, as previously reported by Florvaag and colleagues^12^. However, our study based on a 12-month exposure period was not designed to explore a transient booster effect that might last only a few weeks. This does not preclude that pholcodine would act more on the maturation of B-cell response, increasing the sIgE affinity for QAs rather than on the quantity of IgE produced^8^.

In our study, QA-sIgE and PHO-sIgE had an excellent negative predictive value supporting a low risk of NMBA-related POA when their plasmatic levels are undetectable. This could theoretically help to rule out the risk of NMBA-related POA in patients reporting recent exposure to pholcodine within 12 months. However, this situation may be overwhelming given the common use of pholcodine-containing cough syrups. The value of this strategy would warrant a medico-economic study. In addition, given the low prevalence of NMBA-related anaphylaxis in the general population, estimated at 184 reactions for 1,000,000 anesthesia^1^, and the poor positive predictive value of QA-sIgE and PHO-sIgE, the risk of excessively ruling out NMBAs in past-exposed to pholcodine patients with positive QA-sIgE or PHO-sIgE could exceed the benefits in terms of allergic risk. Therefore, this strategy should not be recommended based only on the ALPHO study results.

## Limitation of the study

Our assessment of pholcodine exposure was based on two imperfect but complementary sources. Regarding drug prescription history retrieved from their community pharmacists, we observed a higher proportion of missing information in controls, possibly related to a weaker motivation of these patients who did not experience NMBA-related POA to collect the required information.

We also observed a weak agreement between the results of the self-administered questionnaires and the drug prescription histories, primarily due to positive answers in the self-questionnaire, not confirmed by the medication history of the pharmacist(s). This can be because patients could report the intake of pholcodine syrups available in their “family medecine cabinet” possibly prescribed or bought for another family member. But interestingly, pholcodine exposure remained significantly associated with NMBA-related POA, whatever the combination of the sources, thus lowering the impact of this weak agreement.

## CONCLUSION

Our study confirms a significant association between pholcodine consumption in the year preceding NMBA exposure and NMBA-related POA. Other environmental factors, including occupational exposure to QA, should be considered in the risk of NMBA-related anaphylaxis, but they currently remain poorly defined. In this context, pholcodine appears to be a well-identified risk factor that can be addressed. These results support the careful re-evaluation of the benefit/risk ratio of pholcodine-containing cough syrups.

The explanatory mechanism for the role of pholcodine remains elusive, but our QA-sIgE and PHO-sIgE investigations support common epitopes between pholcodine and NMBAs. Specific IgE assays for QA and pholcodine have a good negative predictive value but a low positive predictive value. Considering the low prevalence of NMBA-related POA and the potentially large number of subjects exposed to pholcodine, the value of QA-sIgE or PHO-sIgE in assessing the risk of NMBA-related POA resulting from pholcodine exposure cannot be assumed and requires further evaluation.

## Data Availability

All data produced in the present study are available upon reasonable request to the authors

## Acknowledgments

We are grateful to all the members of the ALPHO study group for their commitment all along the study. We sincerely thank Mrs Julie Lecomte (clinical research coordinator) and Mrs Isabelle Adam (data manager) for their involvement in the collection and quality of the data.

