## Supplemental Table S1, Table S2, Table S3, Table S4, Figure S5 for "Pholcodine consumption increases the risk of perioperative anaphylaxis to neuromuscular blocking agents: the ALPHO case-control study"

**Table S1:** **Matching variables between case and control patients.**

|  | Controls  N=334 | Cases  N=167 | p |
| --- | --- | --- | --- |
| **Region** |  |  | 1.00 |
| North | 230 (69) | 115 (69) |  |
| South | 104 (31) | 52 (31) |  |
| **Sex** |  |  | 1.00 |
| Female | 184 (55) | 92 (55) |  |
| Male | 150 (45) | 75 (45) |  |
| **Age** |  |  | 1.00 |
| ≤ 65 years old | 242 (73) | 121 (73) |  |
| > 65 years old | 92 (27) | 46 (27) |  |
| **NMBAs*** |  |  |  |
| Suxamethonium | 210 (63) | 105 (63) | 1.00 |
| Benzylisoquinoline | 148 (44) | 74 (44) | 1.00 |
| Steroidal | 42 (13) | 21 (13) | 1.00 |
| **Season** |  |  | 1.00 |
| Spring/Summer | 144 (43) | 72 (43) |  |
| Fall/Winter | 190 (57) | 95 (57) |  |

NMBA: Neuromuscular blocking agents. Results are expressed as N (%).

* The sum of each NMBA is greater than the patient total because some patients were exposed to multiple NMBAs during the anaesthesia index.

**Table S2:** Type of surgical procedure at the time of patient enrolment.

|  | **Controls**  **N=334** | **Cases**  **N=167** |
| --- | --- | --- |
| Digestive | 108 (32) | 60 (36) |
| Endoscopy | 38 (11) | 8 (5) |
| Orthopaedic | 37 (11) | 22 (13) |
| Ear, Nose and Throat | 32 (10) | 6 (4) |
| Urological | 30 (9) | 15 (9) |
| Bariatric | 28 (8) | 20 (12) |
| Vascular | 18 (5) | 15 (9) |
| Gynaecological | 16 (5) | 10 (6) |
| Thoracic | 14 (4) | 6 (4) |
| Other | 13 (4) | 5 (3) |

Bariatric surgeries were excluded from digestive surgery to be displayed separately.

**Table S3:** Anaesthetic protocol used during the surgery in cases and controls and clinical signs of the perioperative hypersensitivity reaction in cases.

|  | **Cases**  **N=167** | **Controls**  **N=334** |
| --- | --- | --- |
| **Anaesthetic protocol** |  |  |
| Hypnotics |  |  |
| Propofol | 156 (94) | 314 (94) |
| Etomidate | 11 (7) | 17 (5) |
| Ketamine | 62 (37) | 118 (35) |
| Midazolam | 28 (17) | 15 (4) |
| Thiopental | 1 (1) | 3 (1) |
| Opioids |  |  |
| Sufentanil | 102 (61) | 177 (53) |
| Remifentanil | 31 (19) | 98 (29) |
| Alfentanil | 7 (4) | 12 (4) |
| Neuromuscular blocking agents |  |  |
| Atracurium | 53 (32) | 114 (34) |
| Cisatracurium | 21 (13) | 37 (11) |
| Mivacurium | 3 (2) | 1 (0) |
| Vecuronium | .. | .. |
| Rocuronium | 21 (13) | 42 (13) |
| Suxamethonium | 105 (63) | 210 (63) |
| **Clinical signs** |  |  |
| Cardiovascular |  |  |
| Mild hypotension | 13 (8) | .. |
| Cardiovascular collapse | 134 (80) | .. |
| Tachycardia | 65 (39) | .. |
| Bradycardia | 7 (4) | .. |
| Low EtCO_2_ | 15 (9) | .. |
| Arrhythmia | 4 (2) | .. |
| Cardiac arrest | 13 (8) | .. |
| Respiratory |  |  |
| Hypoxemia | 22 (13) | .. |
| Bronchospasm | 71 (43) | .. |
| Cutaneous |  |  |
| Erythema | 86 (51) | .. |
| Urticaria | 17 (10) | .. |
| Oedema | 3 (2) | .. |

Results are expressed as N (%). EtCO_2_: end-tidal CO_2_ level

**Table S4: Factors associated with neuromuscular blocking agents-related perioperative hypersensitivity reactions in case of pholcodine consumption in the matched population.**

|  | **Bivariate analysis** | | **Multivariate analysis** | |
| --- | --- | --- | --- | --- |
|  | **OR** | **p** | **OR** | **p** |
| **PHO-C261 > 0.1 kUI/L** | 67.9 (29.1-158.6) | <0.0001 | 60.7 (25.6-143.8) | <0.0001 |
| **Type of NMBA** |  | 0.0485 |  |  |
| Benzylisoquinolines | 1 |  |  |  |
| Suxamethonium | 2.1 (1.1-4.0) |  |  |  |
| Steroidal | 2.7 (1.1-6.5) |  |  |  |
| **Psychiatric history** | 3.6 (1.4-9.2 | 0.0078 |  |  |
| **QA occupational exposure** | 2.6 (1.1-5.8) | 0.0223 |  |  |
| **Allergic/atopic history** | 1.8 (1.0-3.4) | 0.0552 |  |  |
| **Chronic medication with ARB** | 1.9 (0.9-3.9) | 0.1013 |  |  |

Results of Odds Ratio (OR) are expressed as OR (95% confidence interval); ARB, angiotensin receptor blockers; QA, quaternary ammoniums; PHO-C261, ImmunoCAP® (Uppsala, Sweden) allergens c261 specific IgE.

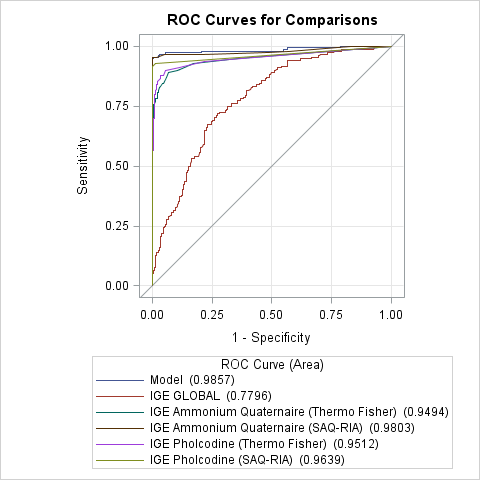

**Figure S5:** ROC curve analysis of specific IgE for quaternary ammonium (QA) and pholcodine (PHO) using the SAQ-FIA or the Immunocap® method (c260, c261) between cases and controls.
